# “We don’t complain; it’s just part of being a woman”: frequency, knowledge, and sociocultural beliefs about dysmenorrhoea in a South African university cohort

**DOI:** 10.64898/2026.06.10.26355353

**Authors:** Gillian J Bedwell, Victoria J Madden, Aaliyah Isaacs, Hlamulo Khorommbi, Nonhlanhla Moloi, Gabriella Papaioannou, Sumaya Solomons, Saniya Sudan, Romy Parker

**Affiliations:** African Pain Research Initiative, Department of Anaesthesia and Perioperative Medicine, Neuroscience Institute, University of Cape Town, Cape Town, Western Cape, South Africa; HIV Mental Health Research Unit, Department of Psychiatry and Mental Health, Neuroscience Institute, University of Cape Town, Cape Town, Western Cape, South Africa; Division of Physiotherapy, Department of Health and Rehabilitation Sciences, University of Cape Town, Cape Town, Western Cape, South Africa

**Keywords:** Dysmenorrhoea, menstrual pain, prevalence, qualitative

## Abstract

**Objective:** Dysmenorrhoea is highly prevalent and affects participation and quality of life. However, little is known about dysmenorrhoea-related knowledge and sociocultural beliefs in South Africa. We investigated dysmenorrhoea frequency, knowledge differences between menstruating and non-menstruating individuals, and associated generational, cultural, and religious beliefs in a South African university cohort.

**Methods:** We analysed cross-sectional survey data from university staff and students. Participants completed demographic and dysmenorrhoea-related knowledge questions and an adapted Working Ability, Location, Intensity, Days of Pain, Dysmenorrhoea (WaLIDD) questionnaire. Free-text responses exploring generational, cultural, and religious beliefs were analysed using reflexive thematic analysis.

**Results:** Of 863 participants, 578 were current or past menstruators. Dysmenorrhoea frequency was 75.4% (95%CI 71.7–78.9), with 53% reporting moderate and 31% severe dysmenorrhoea symptoms. Knowledge of dysmenorrhoea was higher among participants who had menstruated than those who had never menstruated (80.4% vs 55.3%, p<0.001). Most participants (85.1%) wanted more dysmenorrhoea education. Analysis of 246 free-text responses identified five themes: (1) menstrual pain is normalised, dismissed, and expected to endure, (2) reproductive meanings attached to menstrual pain, (3) moral, spiritual, and cultural interpretations of menstrual pain, (4) negotiating competing explanations for menstrual pain, and (5) managing and controlling menstrual pain symptoms. These themes reflected intersecting sociocultural and biomedical understandings of menstrual pain.

**Conclusion:** Dysmenorrhoea was common and symptoms were frequently moderate or severe. Sociocultural beliefs shaped how menstrual pain was understood, communicated, legitimised, and managed. Findings highlight the need to integrate sociocultural perspectives into menstrual health education, support, and healthcare services.

## Introduction

Dysmenorrhoea – pain associated with menstruation – is highly prevalent and interferes with education and work, social participation, physical activity, and quality of life^1–4^. Primary dysmenorrhoea occurs in the absence of identifiable pelvic pathology, whereas secondary dysmenorrhoea is associated with pelvic pathology such as endometriosis^5^. In primary dysmenorrhoea, pain typically begins shortly before or at the onset of menstruation and lasts between 8 and 72 hours^5^. Dysmenorrhoea may also be accompanied by systemic symptoms including gastrointestinal discomfort, sleep and mood disturbances^2^. A recent meta-analysis estimated the global prevalence of dysmenorrhoea to be 71.3% [95%CI: 68.7 – 73.8%]^1^. Epidemiological data from South Africa remain limited. While frequency estimates vary across studies, available evidence suggest that dysmenorrhoea affects between 61% and 77% of South African adolescents and young adults, indicating a burden comparable to global estimates^6,7^. However, prevalence estimates alone provide limited insight into how dysmenorrhoea is experienced and managed. Individuals differ in how they interpret, respond to, and cope with dysmenorrhoea^8–10^. These differences are shaped not only by biological factors, but also by knowledge, social attitudes, and sociocultural beliefs surrounding menstruation and menstrual pain^8,11^.

Consequently, both the experiences and management of dysmenorrhoea are shaped by knowledge, social attitudes, and sociocultural beliefs. In many contexts, menstrual pain is framed as an expected or unavoidable aspect of menstruation and womanhood, reinforcing the idea that dysmenorrhoea should simply be endured rather than treated^8,10,12^. Qualitative studies have demonstrated that beliefs surrounding menstrual pain influence how symptoms are interpreted, whether pain is considered a legitimate health concern, and the extent to which individuals disclose symptoms or seek support. For example, qualitative work from Tanzania found that menstrual pain management was influenced by beliefs that medication could harm the body or impair fertility^12^. Similarly, studies exploring healthcare-seeking behaviours have found that many individuals do not seek care because they perceive dysmenorrhoea as “normal” or anticipate that their pain will be dismissed^9,10^. These beliefs may contribute to delayed healthcare-seeking, reduced use of effective treatments, concealment of symptoms, and ongoing disruption to education, work, and daily activities. Consistent with Good’s argument that diagnostic terms are embedded in sociocultural meaning^13,14^, these findings suggest that dysmenorrhoea is not experienced or managed as a purely biological phenomenon, but as a condition that carries broader social and cultural significance.

Although the influence of sociocultural beliefs on dysmenorrhoea has been explored in several contexts, relatively little research has examined these beliefs within South Africa. South Africa is characterised by considerable cultural, linguistic, and religious diversity and existing South African literature reports that indigenous and cultural understandings influence how menstrual pain is understood and managed^11^. Furthermore, research examining dysmenorrhoea-related knowledge has focused predominantly on menstruating individuals, while the perspectives of non-menstruating individuals remain relatively underexplored despite their important role in shaping social attitudes, support systems, and responses to menstrual health. A better understanding of dysmenorrhoea-related knowledge and beliefs among both menstruating and non-menstruating individuals may provide insight into the sociocultural meanings attached to dysmenorrhoea and how these meanings shape symptom interpretation, disclosure, healthcare-seeking, and treatment utilisation.

Therefore, this study drew on data collected as part of a student-led research project in a South African university community and aimed to (1) determine the frequency of dysmenorrhoea, (2) assess dysmenorrhoea-related knowledge and compare knowledge between menstruating and non-menstruating individuals, and (3) use thematic analysis to explore commonly held generational, cultural, and religious beliefs about dysmenorrhoea in this cohort.

## Methods

### Research design and setting

This analysis uses data collected in a descriptive cross-sectional study examining the frequency, knowledge, and impact of dysmenorrhoea in students and staff of the University of Cape Town (UCT) in 2025. The study protocol was approved by the Faculty of Health Sciences Human Research Ethics Committee (Ref: 862/2024). The project was conducted across all six campuses of UCT, a public university in Cape Town, South Africa, that comprises approximately 28 500 students and 4 650 staff^15^.

### Participants

All consenting adults (≥ 18 years old) who were registered students or staff (full- or part-time) at UCT during data collection were eligible to participate. The study aimed to recruit both menstruating and non-menstruating individuals.

#### Sample Size

Based on a total population of 33 150 staff and students, the sample size was calculated using Yamane’s formula for cross-sectional studies^16^ [n=N/(1+N(e)^2^)], where *n* is the required sample size, *N* is the population size, and *e* is the desired margin of error. Assuming a population size of 33 150 and a margin of error of 5%, the target sample size was 396 participants.

#### Recruitment

Participants were recruited via email, WhatsApp groups, and UCT-affiliated social media platforms. Interested volunteers accessed an online participant information sheet, informed consent form, and survey on the REDCap electronic data capture tool hosted at the University of Cape Town^17,18^. Survey responses were anonymised, REDCap’s response-limiting feature was used to restrict submission to one response per email address. The survey was available in English, Afrikaans, and isiXhosa. Participation was voluntary. Participants could optionally enter a draw to win a ZAR500 (∼USD30) for a large, local online store voucher as an incentive for participation. The survey was available for four weeks during July and August 2025.

### Aims and outcome measures

This study aimed to (1) determine the frequency of dysmenorrhoea within a South African university cohort, (2) assess dysmenorrhoea-related knowledge and compare knowledge between menstruating and non-menstruating individuals, and (3) explore commonly held generational, cultural, and religious beliefs related to dysmenorrhoea using thematic analysis.

All participants completed a demographic questionnaire, which captured their age, sex, home language, religious background, where they grew up (rural vs urban), and their menstruation status (menstruator vs non-menstruator).

#### Aim 1: Determine the frequency of dysmenorrhoea

All participants who had ever menstruated were asked whether they were experiencing or had previously experienced dysmenorrhoea, defined for the purpose of this study as “pain during menstruation that restricts you from participating in daily activities”. Participants also completed an adapted version of the Working ability, Location, Intensity, Days of pain, Dysmenorrhoea (WaLIDD) scale^19^, which combines items on working ability, pain location, pain intensity, and pain duration to estimate the severity of dysmenorrhoea. To provide a more detailed description of pain intensity, the original intensity item was expanded to capture average, least, and worst pain severity experienced during menstruation. Each domain is scored on a four-point ordinal scale (0–3), with higher scores indicating greater severity. Domain scores were summed to generate a total score ranging from 0 to 12, which was categorised as no dysmenorrhoea (0), mild dysmenorrhoea (1–4), moderate dysmenorrhoea (5–7), and severe dysmenorrhoea (8–12) ^19^.

#### Aim 2: Assess dysmenorrhoea-related knowledge and compare knowledge between menstruating and non-menstruating individuals

Both menstruating and non-menstruating participants were asked whether they were familiar with dysmenorrhoea (definition provided), as well as when and how they first learned about it.

#### Aim 3: Explore generational, cultural, and religious beliefs about dysmenorrhoea using thematic analysis

Both menstruating and non-menstruating participants were asked a single question about generational, cultural, or religious beliefs they had been taught about dysmenorrhoea. Responses were entered as free text within the REDCap questionnaire. Participants were also asked whether they personally believed the teachings they had explained in their free text.

### Statistical analysis

Descriptive statistics were used to summarise the data. Normally distributed continuous variables are reported as mean (standard deviation [SD]), while non-normally distributed continuous variables are reported as median (interquartile range [IQR]). Categorical variables are presented as frequencies and percentages.

For Aim 1, the frequency of dysmenorrhoea was calculated and reported with 95% confidence intervals (95%CI). Dysmenorrhoea severity, as measured using the WaLIDD scale, was summarised descriptively across severity categories.

For Aim 2, dysmenorrhoea-related knowledge variables were summarised descriptively for menstruating and non-menstruating participants. Between-group differences in categorical variables were assessed using Pearson’s chi-squared tests or Fisher’s exact tests, where appropriate. Statistical significance was set at p < 0.05.

For Aim 3, responses to the open-ended question exploring generational, cultural, and religious beliefs related to dysmenorrhoea were analysed using reflexive thematic analysis^20,21^. Free-text responses were exported from REDCap into Microsoft Excel for analysis. Data familiarisation was achieved through repeated reading of participant responses. Initial codes were generated inductively from the dataset and grouped into broader code clusters based on conceptual similarity. Candidate themes were identified through iterative review and refinement of these code clusters, with ongoing discussion among the research team to ensure that themes accurately reflected patterns of meaning within the dataset. Themes were subsequently reviewed, defined, and named prior to final analysis and reporting. Quotes are attributed using participant identifiers. Where relevant to interpretation, age range and whether the participant had ever menstruated is indicated (i.e. menstruated vs never menstruated).

We reported the quantitative components of this study in accordance with the Strengthening the Reporting of Observational Studies in Epidemiology (STROBE) guidelines for cross-sectional studies^22^ and the qualitative component in accordance with the Standards for Reporting Qualitative Research (SRQR) guidelines^23^.

### Reflexivity and positionality

The qualitative analysis was informed by a critical realist epistemological stance, which recognises dysmenorrhoea as a real embodied biological phenomenon while acknowledging that understandings and experiences of menstrual pain are shaped through social, cultural, religious, and interpersonal contexts. Consistent with international guidelines for reflexive thematic analyses^21^, themes were viewed as interpretive constructions developed through engagement between the researcher and the data, rather than as objective entities awaiting discovery.

The analysis was further informed by a social constructionist orientation that considers meanings attached to menstrual pain to be negotiated through lived experience, family narratives, cultural practices, religious teachings, and interactions with healthcare and educational systems. Particular attention was therefore paid not only to what participants believed about dysmenorrhoea, but also to how those beliefs shaped the meaning attached to menstrual pain and influenced the ways in which pain was understood, discussed, and managed.

The primary analyst (GJB) is a South African clinical physiotherapist and pain researcher whose clinical and research training is grounded largely within Western scientific frameworks of pain. This background provided a particular interpretive lens through which participants’ accounts were read and understood. GJB also has lived experience of dysmenorrhoea, which may have increased sensitivity to narratives relating to the normalisation, dismissal, and endurance of menstrual pain, as well as accounts describing menstrual pain as biblical punishment. The secondary analyst (VJM) is a South African clinical physiotherapist and researcher with lived experience of well managed menstrual pain. Her training is also grounded within Western scientific frameworks, although she is deeply interested in African perspectives on dysmenorrhoea and pain in general. She has led qualitative studies in South Africa and supported thematic analysis and meta-ethnographic syntheses related to pain.

Throughout coding and theme development, reflexive attention was paid to how these experiences, assumptions, and forms of knowledge shaped interpretation of the data, particularly when engaging with cultural, spiritual, and non-biomedical understandings of dysmenorrhoea. Reflexive notes were maintained throughout the analytic process to document emerging thoughts, emotional responses, assumptions, and interpretations elicited by the data. These reflections were used to critically examine how personal and professional experiences may have influenced coding decisions and theme development. Codes and candidate themes were revisited iteratively and discussed with members of the local pain research team. Codes and candidate themes were also presented at the 2026 National PainSA Congress (Johannesburg) to encourage critical reflection on alternative interpretations and ensure that participants’ accounts were understood as meaningful frameworks through which menstrual pain was interpreted, communicated, and managed, rather than evaluated against biomedical explanations of dysmenorrhoea.

## Results

### Sociodemographic characteristics

Table 1 summarises the sociodemographic characteristics of the participants. A total of 863 participants responded to the survey, with a median (IQR) age of 23 years (20–34). Most participants were female 80.7% (n=619) and reported current menstruation (n=557; 72.6%). Among females who do not currently menstruate, the most commonly reported reasons were post-menopausal (n=44) and health-related conditions (n=22). The total number of past and current menstruators was 578 (67%). Most participants were raised in an urban environment (n=617; 80.5%), primarily spoke English at home (n=408; 53.2%), and identified as coming from Christian households (n=533; 69.5%).

**Table 1:** Summary of the sociodemographic characteristics of the participants (n=863).

|  | **N = 863**<br>Data presented as n (%) or median (IQR) |
|---|---|
| Age (years) | 23 (20 – 34) |
| Missing data | 96 |
| Sex |  |
| Male | 139 (17.7) |
| Female | 619 (80.7) |
| Intersex or variations of sex characteristics | 5 (0.7) |
| Prefer not to say | 6 (0.8) |
| A term not listed | 1 (0.1) |
| Missing data | 96 |
| Do you menstruate? |  |
| Yes | 557 (88.7) |
| No<br>Post-menopausal<br>Pre-menses<br>Non-menstruating (e.g. due to a health condition) | 71 (11.3)<br>44 (62.0)<br>5 (7.0)<br>22 (31) |
| Missing data | 96 |
| Area of upbringing |  |
| Urban | 617 (80.5) |
| Rural | 149 (19.5) |
| Missing data | 97 |
| Language |  |
| Afrikaans (Cape) | 57 (7.4) |
| Afrikaans (Standard) | 29 (3.8) |
| English | 408 (53.2) |
| isiNdebele | 5 (0.7) |
| isiXhosa | 105 (13.7) |
| isizulu | 49 (6.4) |
| SA Sign Language | 1 (0.1) |
| Sepedi | 15 (2.0) |
| Sesotho (Southern Sotho) | 24 (3.1) |
| Setswana | 16 (2.1) |
| Siswati | 4 (0.5) |
| Tshivenda | 23 (3.0) |
| Xitsonga | 9 (1.2) |
| Other | 22 (2.9) |
| Missing data | 96 |
| Religion |  |
| Christian | 533 (69.5) |
| Islam | 88 (11.5) |
| No religious affiliation | 82 (10.7) |
| Other (specify) | 44 (5.7) |
| Traditional African religion | 20 (2.6) |
| Missing data | 96 |

### Missing data

Of the 863 survey responses received, 766 (88.8%) contained complete data across all study variables. Missing data were not imputed. Analyses were conducted using available data, and the extent of missingness is reported for each outcome where relevant.

### Aim 1: Determine the frequency and severity of dysmenorrhoea

Among the past and current menstruators, the frequency [95%CI] of dysmenorrhoea was 75.4% [95%CI 71.7 – 78.9]. Using the WaLLID scale, most participants were categorised as having moderate dysmenorrhoea (n=304; 53%) and 31% (n=176) had severe dysmenorrhoea (Table 2).

**Table 2:**
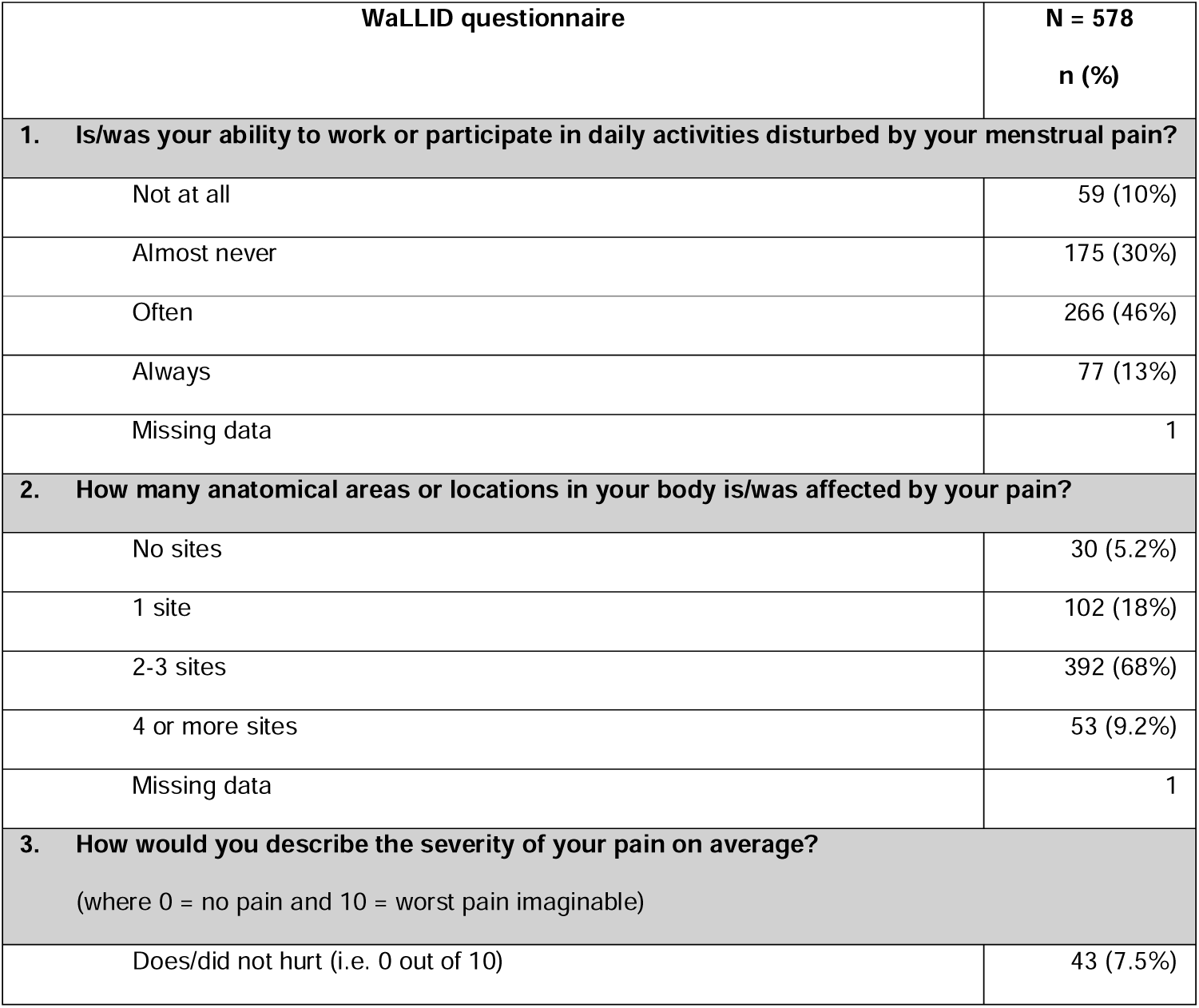
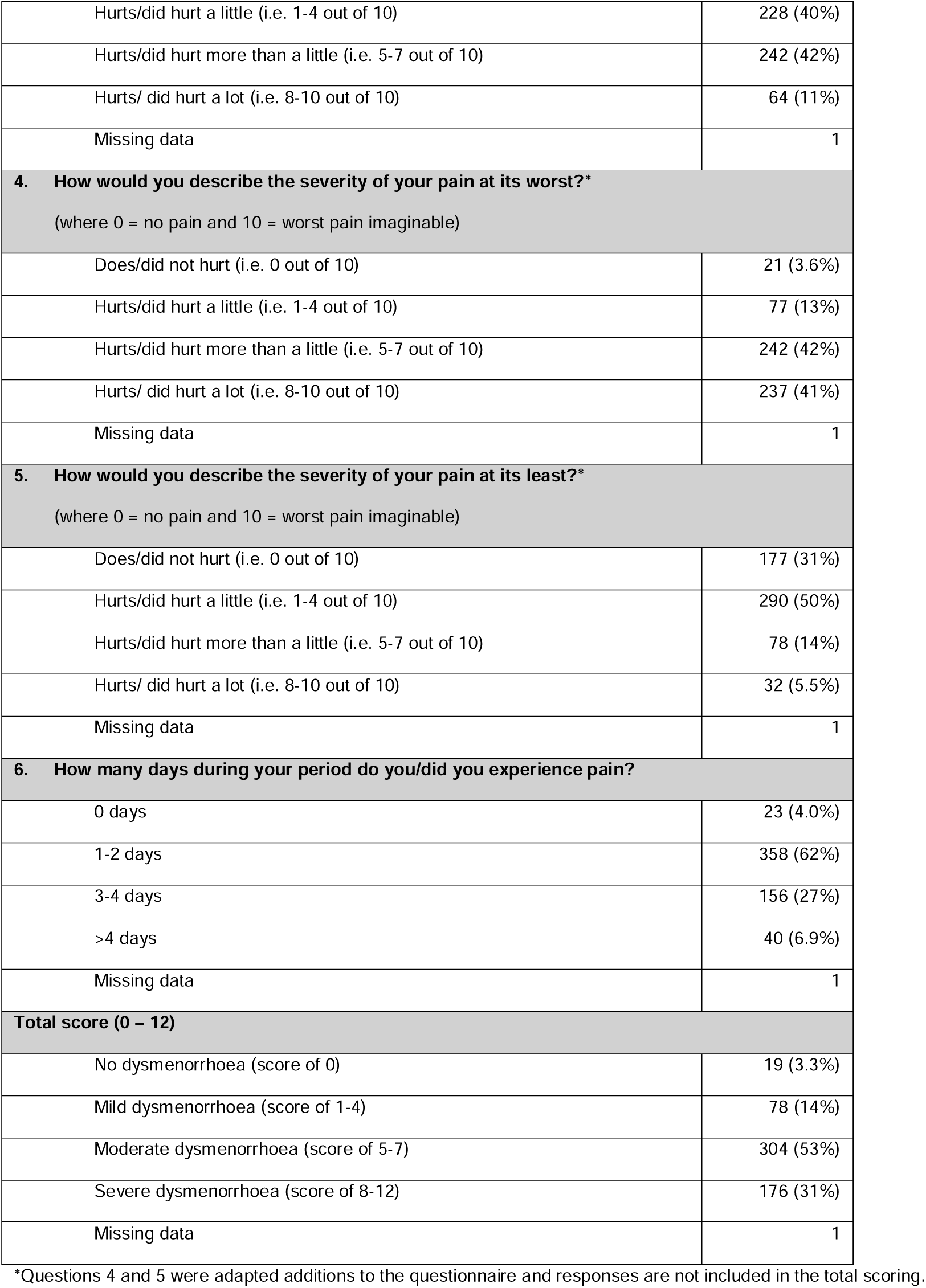
Summary of the data for the WaLLID scale.

| WaLLID questionnaire | N = 578<br>n (%) |
|---|---|
| **1. Is/was your ability to work or participate in daily activities disturbed by your menstrual pain?** |  |
| Not at all | 59 (10%) |
| Almost never | 175 (30%) |
| Often | 266 (46%) |
| Always | 77 (13%) |
| Missing data | 1 |
| **2. How many anatomical areas or locations in your body is/was affected by your pain?** |  |
| No sites | 30 (5.2%) |
| 1 site | 102 (18%) |
| 2-3 sites | 392 (68%) |
| 4 or more sites | 53 (9.2%) |
| Missing data | 1 |
| **3. How would you describe the severity of your pain on average?**<br>(where 0 = no pain and 10 = worst pain imaginable) |  |
| Does/did not hurt (i.e. 0 out of 10) | 43 (7.5%) |
| Hurts/did hurt a little (i.e. 1-4 out of 10) | 228 (40%) |
| Hurts/did hurt more than a little (i.e. 5-7 out of 10) | 242 (42%) |
| Hurts/ did hurt a lot (i.e. 8-10 out of 10) | 64 (11%) |
| Missing data | 1 |
| **4. How would you describe the severity of your pain at its worst?<sup>\*</sup>**<br>(where 0 = no pain and 10 = worst pain imaginable) |  |
| Does/did not hurt (i.e. 0 out of 10) | 21 (3.6%) |
| Hurts/did hurt a little (i.e. 1-4 out of 10) | 77 (13%) |
| Hurts/did hurt more than a little (i.e. 5-7 out of 10) | 242 (42%) |
| Hurts/ did hurt a lot (i.e. 8-10 out of 10) | 237 (41%) |
| Missing data | 1 |
| **5. How would you describe the severity of your pain at its least?<sup>\*</sup>**<br>(where 0 = no pain and 10 = worst pain imaginable) |  |
| Does/did not hurt (i.e. 0 out of 10) | 177 (31%) |
| Hurts/did hurt a little (i.e. 1-4 out of 10) | 290 (50%) |
| Hurts/did hurt more than a little (i.e. 5-7 out of 10) | 78 (14%) |
| Hurts/ did hurt a lot (i.e. 8-10 out of 10) | 32 (5.5%) |
| Missing data | 1 |
| **6. How many days during your period do you/did you experience pain?** |  |
| 0 days | 23 (4.0%) |
| 1-2 days | 358 (62%) |
| 3-4 days | 156 (27%) |
| >4 days | 40 (6.9%) |
| Missing data | 1 |
| **Total score (0 – 12)** |  |
| No dysmenorrhoea (score of 0) | 19 (3.3%) |
| Mild dysmenorrhoea (score of 1-4) | 78 (14%) |
| Moderate dysmenorrhoea (score of 5-7) | 304 (53%) |
| Severe dysmenorrhoea (score of 8-12) | 176 (31%) |
| Missing data | 1 |

<sup>\*</sup>Questions 4 and 5 were adapted additions to the questionnaire and responses are not included in the total scoring.

### Aim 2: Assess dysmenorrhoea-related knowledge and compare knowledge between menstruating and non-menstruating individuals

Table 3 summarises participants’ knowledge of dysmenorrhoea and compares responses between individuals who had ever menstruated and those who had not. Overall, 75.5% (n = 543) of participants reported knowing what dysmenorrhoea is. More individuals who had menstruated reported awareness of dysmenorrhoea than individuals who had never menstruated (80.4% vs 55.3%, p<0.001). The timing of first exposure to the concept of dysmenorrhoea also differed between these groups (p<0.001). Among participants who had heard of dysmenorrhoea, the most commonly reported first sources of information were school or university (32.4%), family (30.7%), friends (23.1%), and media (18.6%). Participants who had menstruated most commonly reported first learning about dysmenorrhoea from family members (33.6% vs 19.1%, p<0.001) or school/university (33.0% vs 29.8%, p=0.46). Among participants who had never menstruated, after school/university sources, the second most common source of information about dysmenorrhoea was friends (26.2% vs 22.3%, p=0.32). Participants who had ever menstruated were more likely than those who had never menstruated to consider menstrual pain that restricts daily activities to be abnormal (55.4% vs 34.8% p<0.001). Most participants (87.5%) denied that speaking about menstruation makes them uncomfortable, and most (84.8%) denied that menstruation was a taboo or stigmatised topic for them. Most participants (85.1%) felt that they should have received more education about dysmenorrhea and its impact, and similar proportions were observed among participants who had menstruated and those who had never menstruated (86.7% vs 78.7%, p=0.05).

**Table 3:**
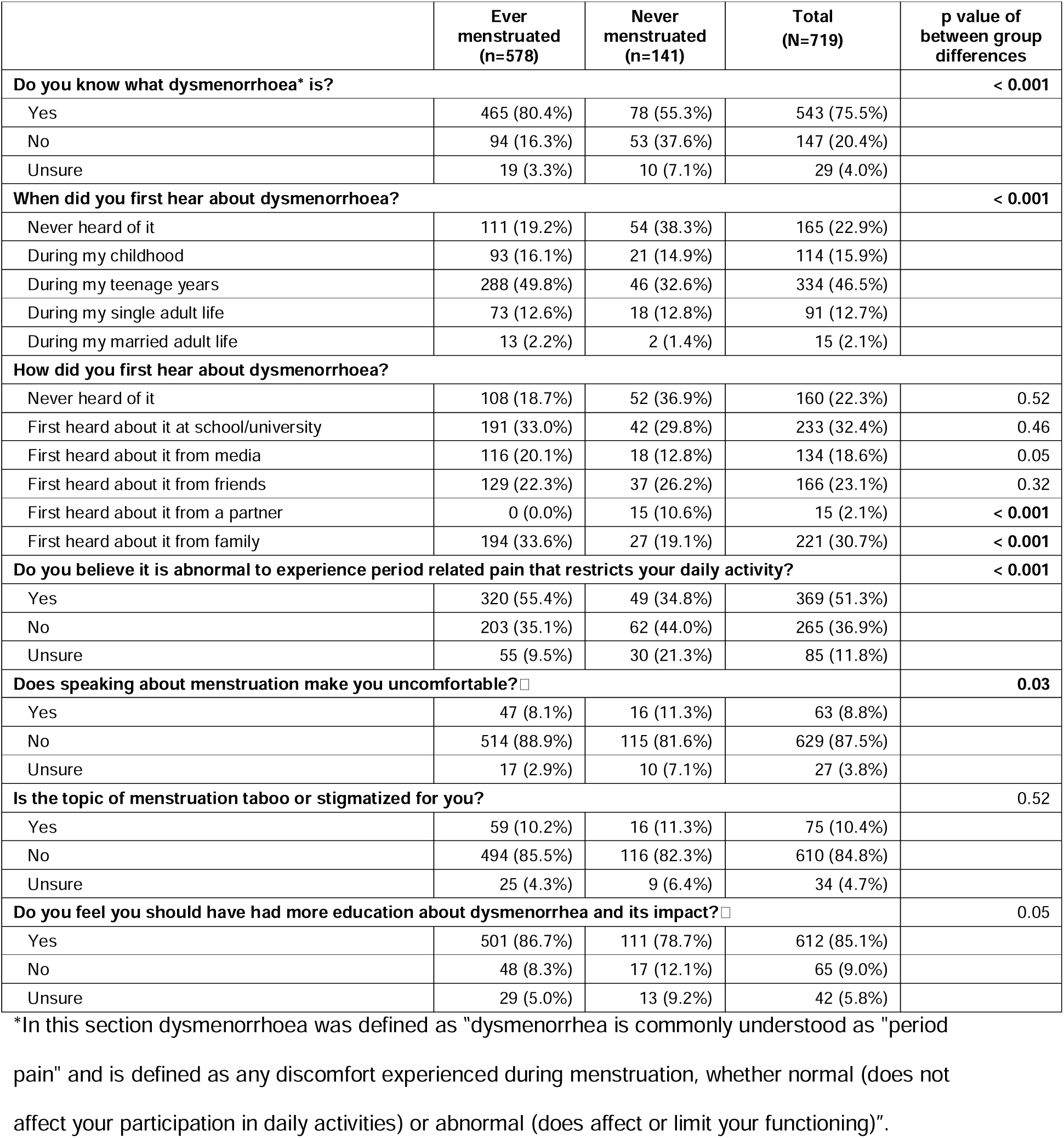
Knowledge of dysmenorrhoea.

|  | **Ever menstruated (n=578)** | **Never menstruated (n=141)** | **Total (N=719)** | **p value of between group differences** |
|---|---|---|---|---|
| **Do you know what dysmenorrhoea\* is?** |  |  |  | **< 0.001** |
| Yes | 465 (80.4%) | 78 (55.3%) | 543 (75.5%) |  |
| No | 94 (16.3%) | 53 (37.6%) | 147 (20.4%) |  |
| Unsure | 19 (3.3%) | 10 (7.1%) | 29 (4.0%) |  |
| **When did you first hear about dysmenorrhoea?** |  |  |  | **< 0.001** |
| Never heard of it | 111 (19.2%) | 54 (38.3%) | 165 (22.9%) |  |
| During my childhood | 93 (16.1%) | 21 (14.9%) | 114 (15.9%) |  |
| During my teenage years | 288 (49.8%) | 46 (32.6%) | 334 (46.5%) |  |
| During my single adult life | 73 (12.6%) | 18 (12.8%) | 91 (12.7%) |  |
| During my married adult life | 13 (2.2%) | 2 (1.4%) | 15 (2.1%) |  |
| **How did you first hear about dysmenorrhoea?** |  |  |  |  |
| Never heard of it | 108 (18.7%) | 52 (36.9%) | 160 (22.3%) | 0.52 |
| First heard about it at school/university | 191 (33.0%) | 42 (29.8%) | 233 (32.4%) | 0.46 |
| First heard about it from media | 116 (20.1%) | 18 (12.8%) | 134 (18.6%) | 0.05 |
| First heard about it from friends | 129 (22.3%) | 37 (26.2%) | 166 (23.1%) | 0.32 |
| First heard about it from a partner | 0 (0.0%) | 15 (10.6%) | 15 (2.1%) | **< 0.001** |
| First heard about it from family | 194 (33.6%) | 27 (19.1%) | 221 (30.7%) | **< 0.001** |
| **Do you believe it is abnormal to experience period related pain that restricts your daily activity?** |  |  |  | **< 0.001** |
| Yes | 320 (55.4%) | 49 (34.8%) | 369 (51.3%) |  |
| No | 203 (35.1%) | 62 (44.0%) | 265 (36.9%) |  |
| Unsure | 55 (9.5%) | 30 (21.3%) | 85 (11.8%) |  |
| **Does speaking about menstruation make you uncomfortable?□** |  |  |  | **0.03** |
| Yes | 47 (8.1%) | 16 (11.3%) | 63 (8.8%) |  |
| No | 514 (88.9%) | 115 (81.6%) | 629 (87.5%) |  |
| Unsure | 17 (2.9%) | 10 (7.1%) | 27 (3.8%) |  |
| **Is the topic of menstruation taboo or stigmatized for you?** |  |  |  | 0.52 |
| Yes | 59 (10.2%) | 16 (11.3%) | 75 (10.4%) |  |
| No | 494 (85.5%) | 116 (82.3%) | 610 (84.8%) |  |
| Unsure | 25 (4.3%) | 9 (6.4%) | 34 (4.7%) |  |
| **Do you feel you should have had more education about dysmenorrhea and its impact?□** |  |  |  | 0.05 |
| Yes | 501 (86.7%) | 111 (78.7%) | 612 (85.1%) |  |
| No | 48 (8.3%) | 17 (12.1%) | 65 (9.0%) |  |
| Unsure | 29 (5.0%) | 13 (9.2%) | 42 (5.8%) |  |

\*In this section dysmenorrhoea was defined as ”dysmenorrhea is commonly understood as "period pain" and is defined as any discomfort experienced during menstruation, whether normal (does not affect your participation in daily activities) or abnormal (does affect or limit your functioning)”.

### Aim 3: Explore commonly held generational, cultural, and religious beliefs related to dysmenorrhoea, using thematic analysis

A total of 246 of the 836 participants (29%) provided free-text responses to the question exploring generational, cultural, and religious beliefs surrounding dysmenorrhoea. Reflexive thematic analysis identified 5 themes: (1) Menstrual pain is normalised, expected, and often silenced, (2) Reproductive meanings attached to menstrual pain, (3) Moral, spiritual, and cultural interpretations of dysmenorrhoea, (4) Negotiating competing explanations for menstrual pain, (5) Managing and controlling menstrual pain symptoms (Fig 1).

**Figure 1:**
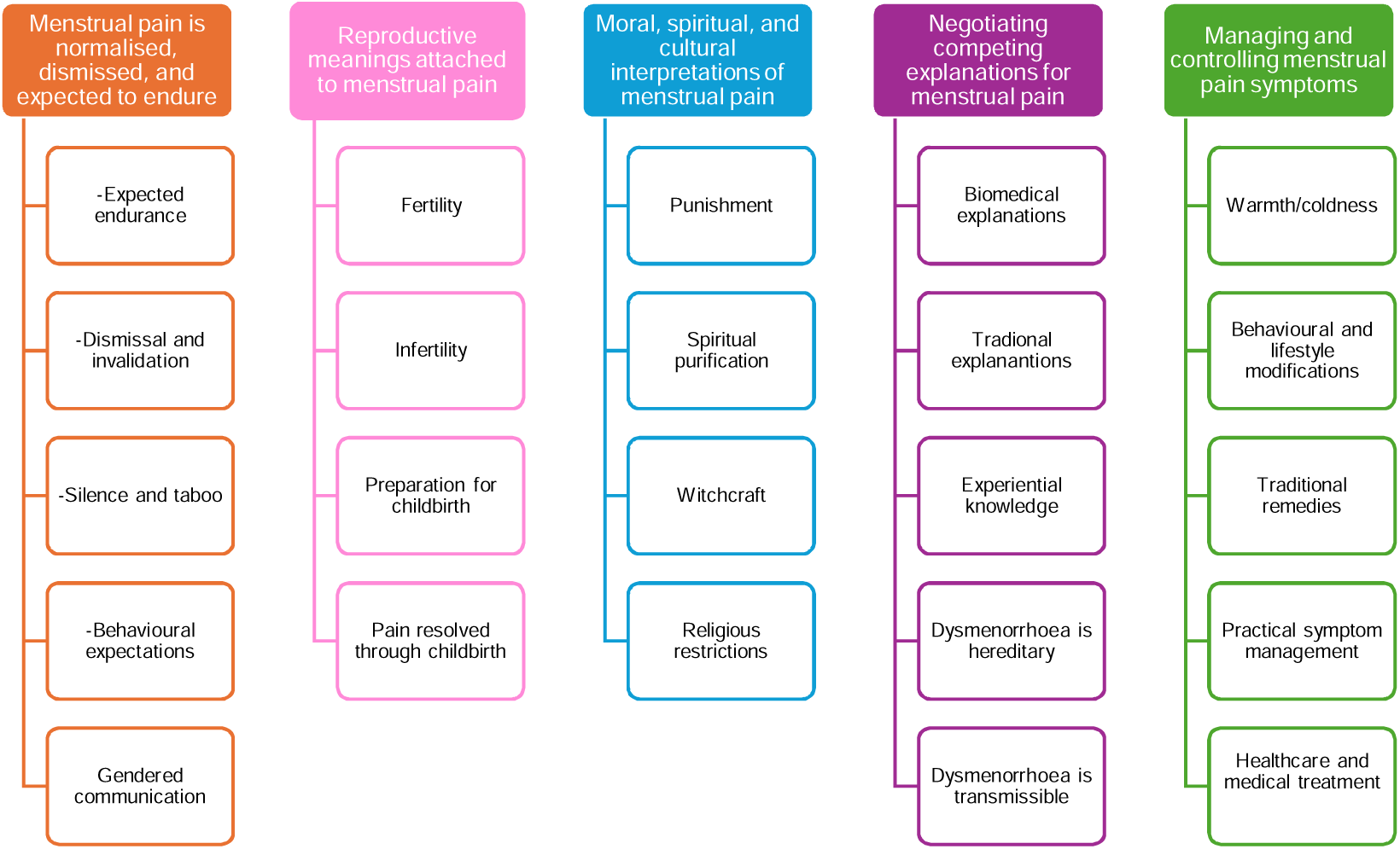
Themes and subthemes identified through reflexive thematic analysis.

#### Theme 1: Menstrual pain is normalised, dismissed, and expected to endure

Participants commonly described dysmenorrhoea as a normalised aspect of menstruation and womanhood. Menstrual pain was framed as an expected and unavoidable experience that women were required to accept rather than address. Participants frequently reported being taught that period pain was “normal”, and that “everyone gets it”. In many accounts, these messages extended beyond recognising pain as common and instead established expectations regarding how pain should – or should not – be expressed. As one participant explained, “We don’t complain. It’s just part of being a woman” (P104: age range 40 – 49y/o, menstruated), while another described being taught that “It’s part of life and woman should endure and suffer in silence” (P46: age range 30 – 39y/o, menstruated).

Importantly, normalisation was frequently accompanied by dismissal and invalidation. Participants described severe or debilitating symptoms being minimised by family members, teachers, and peers, often because pain was perceived as an ordinary part of menstruation. One participant reflected, “Everyone told me it’s normal to have painful periods. Even when I couldn’t walk” (P318: age range 18 – 29y/o, menstruated), while another reported “Other women in my family told me that I was dramatic and faking my pains because theirs wasn’t nearly as painful as mine” (P495: age range 18 –29y/o, menstruated). Several participants critically reflected on the consequences of these beliefs, describing how normalisation delayed recognition of underlying conditions such as endometriosis and discouraged help-seeking: “Invalidating someone’s experience delays the diagnostic process and could even deny them a proper diagnosis” (P273: age range 18 – 29y/o, menstruated). Although less prominent, some participants distinguished between expected menstrual discomfort and severe pathological pain – “Some pain is normal but not severe pain” (P127: age range 60 – 69y/o, menstruated); however, the dominant narrative positioned dysmenorrhoea as something to be endured rather than investigated.

Communication about menstruation and menstrual pain varied across participants’ accounts. Some participants described dysmenorrhoea as a topic that was rarely discussed openly within families and communities, often characterised by silence, secrecy, or stigma. Menstrual pain was framed as a private matter that should be endured rather than openly disclosed, with one participant explaining that “it was something that was not to be spoken about. It was something that women should experience in silence. It was almost a myth in a sense” (P313: age range 18 – 29y/o menstruated). Others similarly explained that it was “not something that was spoken about openly” (P166: age range 30 – 39y/o menstruated) or that “no one talks about ‘lady things’ in the family” (P395: age range 18 – 29y/o menstruated). For some participants, these communication norms were explicitly gendered, with discussions of menstruation restricted to women and excluded from conversations with men: “Culturally we don’t speak about it with male family members” (P574: age range 18 – 29y/o menstruated).

Conversely, some participants described contexts in which menstruation and menstrual pain could be discussed openly. Some reported supportive family environments in which menstruation was treated as a normal topic of conversation, noting that “it was not a taboo subject in our home at all” (P256: age range 50 – 59y/o, menstruated). Others described learning about menstrual pain through schooling or partners. For instance, one participant reflected, “My family preferred not to speak about it, indeed the topic was never brought up in my home. My primary school ran a workshop on sexual education which included information and period pains. My partner and I freely speak about menstruating and period pains which has really helped [me] understand it better. She is the first female close to me who openly shares the details of the discomfort” (P777: age range 18 – 29y/o, never menstruated). These accounts suggest conflicting narratives governing when, where, and with whom menstrual pain could be discussed.

#### Theme 2: Reproductive meanings attached to menstrual pain

Participants frequently interpreted dysmenorrhoea through reproductive and gendered meanings, rather than solely physiological explanations. Participants linked dysmenorrhoea to beliefs about fertility and future childbearing, although these beliefs were often contradictory. For some participants, menstrual pain was framed positively, as a sign of strong fertility or reproductive potential. For instance, one participant reflected, “Culturally, if you experience dysmenorrhea, you are most likely very fertile.” (P148: age range 30 – 39y/o, menstruated), while another reported, “I have been told [that] people with severe period pain will get twins” (P307: age range 18 – 29y/o, menstruated). In some accounts, pain was positioned almost as evidence of reproductive blessing, with one participant describing, “That you must push through it [the pain] since it is a gift that indicates fertility which is a blessing in itself” (P477: age range 18 – 29y/o, menstruated).

In contrast, other participants described beliefs that menstrual pain reflected reproductive problems or threatened future fertility. One participant reflected, “My relatives used to say you won’t be able to bear children if you experience period pains” (P803; age range 30 – 39y/o, menstruated) and another reported “Having period pains can affect your ability to have babies” (P511: age range 18 –29y/o, menstruated). Concerns about infertility also extended to the management of pain itself, particularly around the use of medication. One participant described “Grandmother and aunties from my mother’s side believe that taking period pain medication or just any pain medication will negatively affect pregnancy or cause infertility” (P614: age range 18 – 29y/o, menstruated).

Participants also described menstrual pain as serving a purpose by preparing women for childbirth, with one participant explaining “it [menstrual pain] prepares one for childbirth” (P279: age range 30 – 39y/o, never menstruated) and another described “the pain you feel is a symbol of what is to come when you’re giving birth so period pains help prepare you for that” (P821: age range 18 – 29y/o, menstruated). Childbirth itself was portrayed as both the culmination of menstrual suffering and its resolution, with recurring messages that pain would disappear after pregnancy, childbirth, or sexual activity.

Together, these narratives suggest that dysmenorrhea was interpreted through a reproductive lens, with menstrual pain understood in relation to fertility, childbearing, and reproductive expectations. However, the meaning attached to pain was inconsistent, with dysmenorrhea framed by some as a marker of heightened fertility, and by others as a sign of impaired reproductive potential.

#### Theme 3: Spiritual, moral, and cultural interpretations of menstrual pain

For many participants, dysmenorrhoea was interpreted through broader spiritual, religious, and moral frameworks. Participants linked menstrual pain to religious narratives of female suffering, particularly those associated with the biblical Eve and punishment. Participants described being taught that painful menstruation was a consequence of sin, with one participant explaining “it is normal to have debilitating pain for women because Eve sinned hence this is the punishment” (P320: age range 18 – 29y/o, menstruated). In these accounts, menstrual pain was imbued with moral significance and interpreted as an expected punitive burden associated with womanhood.

Participants described beliefs that menstruation rendered women spiritually impure or unclean, whereas others assigned a positive spiritual meaning to menstrual pain itself, viewing menstrual pain as a process of spiritual purification, for instance one participant reflected “I was taught that period pains are spiritually, they may mean purification which is starting afresh” (P215: age range 18 – 29y/o, menstruated). Across different religious and cultural contexts, menstruation was associated with restrictions on prayer, fasting, cooking, entering places of worship, and participation in social activities. There were also accounts linking severe dysmenorrhoea to witchcraft, curses, or supernatural causes. One participant described, “Culturally, I was taught that having severe period pains like me meant that I had a curse called “selomi”. It is believed that I have something alive in my womb (like a parasite or something like that) and during period pains, it moves around and causes me pain” (P286: age range 18 – 29y/o, menstruated).

There was variation in how these beliefs were experienced and enforced. Some participants noted that beliefs were common in extended family or community settings but less present within their immediate households: “Although it [the belief] was never present in my immediate family, it was rife in my extended family” (P347: age range 18 – 29y/o, never menstruated). Together, these narratives suggest that the meaning of dysmenorrhoea was embedded within broader systems of spiritual, cultural, and moral meaning, which shaped how pain was interpreted, discussed, and managed.

#### Theme 4: Negotiating competing explanations for menstrual pain

Participants described drawing on multiple and sometimes competing systems of knowledge to understand dysmenorrhoea. Explanations ranged from biomedical understandings grounded in anatomy, pathology, and healthcare to traditional, familial, and experiential frameworks. Some participants described learning about menstrual pain through school, healthcare professionals, and family histories of conditions such as endometriosis and polycystic ovarian syndrome. These participants often distinguished expected menstrual discomfort from pain severe enough to warrant medical investigation. As one participant explained, “I grew up understanding that most females get period pain but if they [the pains] are severe to the point that you can’t get out of bed, then that’s abnormal and it’s time to visit the doctor” (P522: age range 18 – 29y/o menstruated).

Biomedical explanations frequently coexisted alongside traditional understandings, rather than replacing them. Participants described beliefs that menstrual pain resulted from inherited vulnerability, transferable influences, or cold exposure. Some participants reported being taught that sharing items of clothing, particularly those that sit on the waistline, e.g. skirts, trousers, belts, or underwear, could transfer dysmenorrhoea between individuals. One participant explained, “My gran said that I developed them [menstrual pain] because I wore something that touched my waist that belonged to someone who had cramps” (P583: age range 18 – 29y/o menstruated), another participant reflected “it [menstrual pain] can be passed from people who suffer from it to those who don’t through sharing clothes especially those that sit on the waist such as pants/skirts” (P669: age range 30 – 39y/o menstruated).

Participants also described beliefs that walking barefoot or sitting on cold surfaces could cause or worsen their menstrual pain, for instance one participant reported being taught “not to be barefoot on cold floors or sit on cold surfaces as doing so will exacerbate your cramps” (P580: age range 18 – 29y/o menstruated). Although some responses simply described cold as a trigger for pain, others embedded this belief within a broader explanatory framework in which dysmenorrhoea was attributed to the presence of blood clots. Within this narrative, menstrual pain was understood to result from retained or stagnant blood clots, while warmth was believed to facilitate their dissolution or expulsion. For example, one participant explained that “Cramping usually means there are blood clots, take half a Disprin [aspirin] to help thin the blood, keep warm”(P203: age range 30 – 39y/o, menstruated). Another described how steaming practices resolve clots and alleviate menstrual pain: “Sit on a bucket filled with hot water, onions and Salvon [*sic* Savlon; antiseptic product] and let the steam go into your vagina and the period blood clots come out, the pain will be better” (P354: age range 18 – 29y/o, menstruated). These narratives suggest that beliefs about cold exposure and warmth were not merely behavioural recommendations but reflected broader understandings of the underlying causes of menstrual pain. Taken together, these accounts illustrate how participants navigated multiple explanatory frameworks simultaneously, often assimilating biomedical, cultural, familial, and experiential knowledge when making sense of menstrual pain.

#### Theme 5: Managing and controlling menstrual pain symptoms

Participants described a wide range of strategies intended to prevent, alleviate, or control dysmenorrhoea. Many management practices centred on maintaining warmth, including the use of hot water bottles, warm drinks and herbal teas, steaming practices, and avoiding sitting on cold surfaces. Participants reported being taught that warmth protected the reproductive system and relieved cramping, whereas cold exposure was believed to worsen symptoms. Dietary practices also featured prominently, with restrictions on foods such as dairy products, eggs, and sugary foods, which were believed to exacerbate menstrual pain. These narratives reflected beliefs that menstrual pain could be influenced through behavioural and lifestyle modifications.

Alongside these approaches, participants described a mixture of biomedical and traditional remedies. Some reported using western pain medication, whereas others described herbal therapies and steaming practices. For instance, one participant described “Dysmenorrhoea can be cured by traditional herbs that clean the womb” (P775: age range 18 – 29y/o, menstruated). Advice regarding management was often contradictory. While some participants were encouraged to use analgesics, others reported being warned that pain medication could impair fertility or future pregnancy. These findings suggest that menstrual pain management was shaped by diverse and sometimes competing understandings of health, reproduction, and the body.

In summary, dysmenorrhoea was rarely understood as “just” a physical symptom. Instead, menstrual pain appeared to function as a socially and culturally interpreted experience through which ideas about womanhood, reproduction, morality, bodily regulation, endurance, and femininity were communicated and reinforced. Across the dataset, dysmenorrhoea was often normalised, moralised, concealed, regulated, or symbolically interpreted in ways that shaped how pain was experienced, expressed, managed, and legitimised. Finally, more individuals who had menstruated denied believing their reported generational, cultural, and religious beliefs surrounding dysmenorrhoea than individuals who had never menstruated (63.2% vs 43.9% p<0.001; Supplementary material, Table S1).

## Discussion

This study drew on data collected as part of a student-led research project in a South African university community and found that three-quarters of those who had menstruated reported dysmenorrhoea. Awareness of dysmenorrhoea differed between menstruating and non-menstruating individuals, and most participants reported wanting more education about dysmenorrhoea and its impact. Reflexive thematic analysis identified five interconnected themes relating to generational, cultural, and religious beliefs surrounding dysmenorrhoea: (1) Menstrual pain is normalised, expected, and often silenced, (2) Reproductive meanings attached to menstrual pain, (3) Moral, spiritual, and cultural interpretations of dysmenorrhoea, (4) Negotiating competing explanations for menstrual pain, and (5) Managing and controlling menstrual pain symptoms. Together, these findings suggest that dysmenorrhoea is rarely understood as a purely biological phenomenon. Rather, dysmenorrhoea is interpreted through social, cultural, reproductive, spiritual, and biomedical frameworks that shape how pain is understood, communicated, and managed.

The frequency of dysmenorrhoea observed in this cohort is broadly comparable to global estimates^1,24^ and supports growing evidence that menstrual pain represents a substantial public health concern globally^6^. While prevalence estimates vary according to how dysmenorrhoea is defined and measured, the symptom burden observed in this cohort was considerable, especially considering our definition for dysmenorrhoea reflected high impact pain: “pain during menstruation that restricts you from participating in daily activities”. Although approximately three-quarters of participants who had menstruated met this definition, only 3.3% of participants were classified as having no dysmenorrhoea on the WaLIDD scale, while more than four-fifths were classified as having moderate (53%) or severe (31%) dysmenorrhoea. These findings suggest that menstrual pain is not only common, but is frequently experienced at levels likely to interfere with daily functioning. Despite this substantial burden, participants commonly described menstrual pain as a normal, expected, and inevitable part of menstruation and womanhood. Narratives of endurance, dismissal, and invalidation were evident across cultural and religious contexts, with participants frequently describing expectations that pain should simply be tolerated rather than recognised as a legitimate health concern. This normalisation may be particularly problematic because it risks obscuring the distinction between expected menstrual discomfort and clinically significant pain, potentially contributing to delayed healthcare-seeking, reduced treatment utilisation, and the continued under-recognition of dysmenorrhoea and underlying pathologies such as endometriosis^25^.

The qualitative findings suggest that dysmenorrhoea functions as more than a physiological symptom; it is also a socially and culturally meaningful experience through which broader ideas about womanhood, reproduction, morality, spirituality, and bodily functioning are communicated and negotiated. Participants described inheriting diverse and sometimes contradictory explanations for menstrual pain, including beliefs linking dysmenorrhoea to fertility, infertility, childbirth, punishment, purification, heredity, transmissible, and spiritual causes. Importantly, these beliefs were not simply accepted or rejected. Rather, participants appeared to actively negotiate, reinterpret, and sometimes resist inherited understandings of menstrual pain. This finding aligns with Good ^14^’s conceptualisation of illness meanings, which proposes that symptoms acquire significance through the social and cultural frameworks used to interpret them. Within this framework, dysmenorrhoea is not experienced solely as pain, but as pain that carries particular meanings, expectations, and social consequences.

Dysmenorrhoea is associated with psychological distress^26^ and poorer mental health outcomes^27^. While the present study did not assess psychological outcomes, narratives around stigma, dismissal, invalidation, and expected endurance were common. There is growing evidence linking stigma with poorer outcomes across chronic pain conditions^28^, and stigmatisation and shaming could be partly responsible for (or exacerbate) mental health consequences of dysmenorrhoea^26–28^.

Taken together, social meanings attached to menstrual pain may contribute to psychological distress and poorer mental health outcomes for some menstruating individuals and warrants future investigation. Dysmenorrhoea has also emerged as a risk factor for later chronic pain^29^. The recurrent anticipatory nature of menstrual pain as been linked with heightened levels of pain-related expectations, fear, and worry^30–32^, which may contribute to chronic pain risk. Therefore, understanding how individuals learn to interpret, anticipate, and respond to menstrual pain may therefore be important for understanding both the immediate and longer-term consequences of dysmenorrhoea.

An interesting tension emerged between the quantitative and qualitative findings. While qualitative narratives described menstrual pain as stigmatised, silenced, or dismissed, most participants reported that speaking about menstruation did not make them uncomfortable and denied that menstruation was a taboo topic. Furthermore, approximately 60% of participants reported that they did not personally believe the generational, cultural, or religious teachings they described. At face value, these findings appear contradictory. However, the qualitative question invited participants to reflect on beliefs and messages they had been taught, whereas the quantitative questions assessed their current attitudes and experiences. As such, the findings may suggest that participants were able to recognise and articulate inherited beliefs without necessarily endorsing them. This interpretation aligns with research suggesting that although menstrual stigma remain pervasive, younger and more educated populations may be increasingly willing to discuss menstruation openly and challenge traditional beliefs^33–35^. Studies from South Africa and other settings have similarly reported tensions between increasing openness about menstruation and the persistence of stigma, secrecy, and expectations of endurance surrounding menstrual pain^36,37^. Although many participants in the present study rejected the beliefs they described, qualitative accounts suggest that these beliefs may continue to shape social expectations regarding disclosure, help-seeking, and symptom management. Social norms do not necessarily require conscious endorsement to exert influence; rather, they may persist through family practices, cultural narratives, and community expectations. The coexistence of apparent openness alongside narratives of silence and stigma therefore likely reflects an ongoing negotiation between inherited understandings of dysmenorrhoea and evolving contemporary attitudes toward menstruation.

### Strengths and limitations

This study has three main strengths. First, it included a large and socio-culturally diverse sample of university staff and students, with participants representing 11 of 12 official South African languages and a range of cultural, religious, and geographic backgrounds. Second, unlike many studies of menstrual health, this study included both menstruating and non-menstruating participants, allowing exploration of dysmenorrhoea-related knowledge beyond those with direct lived experience of menstrual pain. Third, the integration of quantitative and qualitative data provided insight not only into the frequency and severity of dysmenorrhoea, but also into the meanings, beliefs, and social contexts through which menstrual pain is understood and managed.

This study also has limitations. Participants were recruited from a single South African university and consisted primarily of highly educated students and staff. The findings may not be generalisable to individuals with different educational, socioeconomic, or healthcare access profiles, although the inclusion of participants from diverse linguistic, cultural, and religious backgrounds did reveal a range of beliefs about dysmenorrhoea in South Africa.

The qualitative component drew on responses to a single typed survey question. While this approach enabled broad participation and captured a diverse range of perspectives, the resulting data were relatively thin and did not allow for in-depth exploration of participants’ beliefs and experiences. As such, the findings should be interpreted as an exploration of commonly reported beliefs rather than a comprehensive account of how dysmenorrhoea is understood and experienced. Future qualitative research using interviews, focus groups, or community-based participatory approaches is needed to explore the origins, meanings, and implications of these beliefs in greater depth.

## Conclusions

Dysmenorrhoea was common in this South African university cohort and was associated with symptoms suggestive of moderate to severe dysmenorrhoea. While most participants reported awareness of dysmenorrhoea and expressed a desire for greater education about menstrual pain, qualitative findings demonstrated that dysmenorrhoea was rarely understood as a purely biological phenomenon. Instead, menstrual pain was interpreted through intersecting social, cultural, reproductive, spiritual, and biomedical frameworks that shaped how pain was recognised, discussed, legitimised, and managed.

Participants described narratives that normalised menstrual pain, encouraged endurance, and linked dysmenorrhoea to broader meanings surrounding fertility, morality, spirituality, and bodily functioning. At the same time, many participants reported not believing their inherited narratives surrounding menstrual pain, suggesting an ongoing negotiation between traditional understandings of menstrual pain and contemporary perspectives. These findings highlight the importance of addressing dysmenorrhoea not only as a clinical or public health issue, but also as a socially and culturally situated experience.

Improving menstrual health outcomes may therefore require approaches that extend beyond symptom management alone and, instead, engage with the beliefs, meanings, and social contexts through which dysmenorrhoea is understood. Further qualitative research is needed to explore how these beliefs influence healthcare-seeking, treatment utilisation, psychological wellbeing, and longer-term pain outcomes in diverse South African communities.

## Supporting information

STROBE checklist

## Data Availability

All data produced in the present study are available upon reasonable request to the authors.

## Author contributions

AI, HK, NM, GP, SS, SS, and RP conceptualised the study, developed the methodology, and oversaw data collection. GJB led the data analysis, with all authors contributing to the interpretation of findings. GJB prepared the original manuscript draft. All authors critically reviewed and revised the manuscript and approved the final version for publication.

