## Supplementary material: Table S1 for "“We don’t complain; it’s just part of being a woman”: frequency, knowledge, and sociocultural beliefs about dysmenorrhoea in a South African university cohort"

Table S1: Summary of whether participants endorsed or denied believing their reported generational, cultural, and religious beliefs surrounding dysmenorrhoea.

|  | **Ever menstruated (n=578)** | **Never menstruated (n=141)** | **Total**  **(N=719)** | **p value of between group differences** |
| --- | --- | --- | --- | --- |
| **Do you believe your generational/cultural/religious teachings surrounding dysmenorrhoea?** | | | | < 0.001 |
| Yes | 72 (17.3%) | 20 (20.4%) | 92 (17.9%) |  |
| No | 263 (63.2%) | 43 (43.9%) | 306 (59.5%) |  |
| Unsure | 81 (19.5%) | 35 (35.7%) | 116 (22.6%) |  |
| Missing data | 162 | 43 | 205 |  |
